# Low-field thoracic magnetic stimulation increases peripheral oxygen saturation levels in coronavirus disease (COVID-19) patients: a single-blind, sham-controlled, crossover study

**DOI:** 10.1101/2021.05.21.21256456

**Authors:** Saul M Dominguez-Nicolas, Elias Manjarrez

## Abstract

The severe acute respiratory syndrome coronavirus-2 (SARS-CoV-2) may cause low oxygen saturation (SpO2) and respiratory failure in coronavirus disease (COVID-19) patients. Hence the increase of SpO2 levels could be crucial for the quality of life and recovery of these patients. Here we introduce an electromagnetic device termed low-field thoracic magnetic stimulation (LF-ThMS) system. This device was designed to non-invasively deliver a pulsed magnetic field from 100 to 118 Hz and 10.5 to 13.1 mT (i.e., 105 to 131 Gauss) to the dorsal thorax. We show that these frequencies and magnetic flux densities are safe for the patients. We also present a proof-of-concept that a single session of LF-ThMS applied for 30 minutes to the dorsal thorax of 17 COVID-19 patients significantly increases their SpO2 levels. We designed a single-blind, sham-controlled, crossover study on 5 COVID-19 patients who underwent two sessions of the study (real and sham LF-ThMS) and 12 COVID-19 patients who underwent only the real LF-ThMS. We found a statistically significant correlation between magnetic flux density, frequency, or temperature associated with the real LF-ThMS and SpO2 levels in all COVID-19 patients. However, the five patients of the sham-controlled study did not exhibit a significant change in their SpO2 levels during sham stimulation. All the patients did not present adverse events after the LF-ThMS intervention.

**Trial registration:** ClinicalTrials.gov

**Identifier:** NCT04895267

## Introduction

Although recent studies about the structure and function of SARS-CoV-2 may help develop new targeted treatments against COVID-19, there is still not a universally approved treatment for this sickness [1,2]. For instance, some pharmacological treatments include the controversial use of azithromycin, ivermectin, oseltamivir, remedesivir, favipiravir, tocilizumab, ribavirin, lopinavir, interferon β-1b, lopinavir/ritonavir, hydroxychloroquine, or chloroquine phosphate (for review, see [1,3,4]). Many of them are based mainly on case studies, prospective, or retrospective observational studies, with a low number of randomized controlled trials and low quality of study design to guarantee their efficacy and safety [5]. Moreover, in severe cases, many countries employ empiric antimicrobial therapy, mechanical ventilation, convalescent plasma therapy, or combinations of antiviral and anti-inflammatory drugs [6,7]. Because fever and acute respiratory failure are common symptoms, the management of these patients includes antipyretics and oxygen therapy to increase SpO2 levels during respiratory distress. Hence the development of new methods to increase SpO2 levels in COVID-19 patients could become a potential complement of oxygen masks, ventilators, or other modalities to improve oxygenation.

This study aimed to present a proof-of-concept that a 30 minutes single-session of dorsal LF-ThMS can be employed to increase SpO2 levels in COVID-19 patients significantly. We hypothesized that the variables associated with LF-ThMS, as frequency, magnetic flux density, and temperature in the dorsal thorax, might be correlated to SpO2 levels in these patients. Our proof-of-concept research could be helpful to design future randomized controlled trials intended to develop plausible LF-ThMS treatments in the successful management of these patients.

We acknowledge that several magnetic field interventions are controversial, but others are gaining an excellent reputation as the transcranial magnetic stimulation. This controversy could be due to the low quality of study designs alongside the exaggerated promotion of alternative therapies intended only for lucrative practices. Therefore, to avoid misinterpretations with our research, we included a single-blind, sham-controlled, crossover study on 5 COVID-19 patients who underwent two sessions of the study (real and sham LF-ThMS) and 12 COVID-19 patients who underwent only the real LF-ThMS. Moreover, we applied LF-ThMS in a short time range, i.e., in a 30 minutes single-intervention to measure SpO2 values. In this form, we avoided confounding factors related to the spontaneous recovery by natural immunity, common in many COVID-19 patients several days after the contagion.

Regarding safety, our LF-ThMS device applied to the dorsal thorax produces low-intensity magnetic flux densities in a safe range of 10.5 to 13.1 mT at 100 to 118 Hz. Such magnetic fields are within the frequency range of extremely low-frequency (0–300 Hz) magnetic fields (ELF-MFs) to study the interaction between ELF-MFs and neuronal systems [8,9,10]. Our LF-ThMS device also produces heat with a safe temperature range from 27.5 to 44 °C, consistent with the well-known tolerance of the isolated and perfused dog lung to hyperthermia in this temperature range [11,12]. Rickaby et al. (1991) [11] found that temperatures below 44.4 °C for two hours had no detectable influence on the following measured variables of lung weight, extravascular water, vascular volume, serotonin uptake, urea permeability, surface area product, perfusion pressure, and lung compliance. In line with such findings, Cowen et al. (1992) [12] confirmed that the isolated dog lung with perfusion was tolerant to hyperthermia up to about 44 °C for 1 hour. Other studies claim that hyperthermia in this range is beneficial and enhances the immune response [13,14,15].

Our results demonstrate that LF-ThMS locally applied to the dorsal thorax of COVID-19 patients is safe and valuable to significantly increase SpO2 levels during a single LF-ThMS intervention of 30 minutes. This is in line with models predicting the electrical [16] or thermal inactivation of SARS-CoV-2 [17] in the environment or with the hypothesis that hydro-thermotherapy or photobiomodulation could help in the treatment of COVID-19 patients [18,19].

## Materials and methods

### Study design

We designed a single-blind, sham-controlled, crossover study on 5 COVID-19 subjects who underwent two sessions of the study (sham or real LF-ThMS) and 12 COVID-19 subjects who underwent only the real LF-ThMS stimulation. The study was performed following the Declaration of Helsinki and approved by a local ethics committee from the Benemérita Universidad Autónoma de Puebla, Mexico (protocol: Oficio No. SIEP/C.I./065A/2020, book number: 2, sheet number: 133, registration number: 818, date: July 3, 2020). The study was registered in ClinicalTrials.gov (Identifier: NCT04895267). All subjects voluntarily participated with full understanding and signed informed consent. The SPIRIT-Checklist is included as a supplementary file.

### Patients

We applied LF-ThMS on the dorsal thorax to 17 patients (25-81 years of age) who were selected according to the following criteria. The inclusion criteria were: 1) Adult patients diagnosed with mild to moderate COVID-19 disease without pneumonia. The COVID-19 disease severity was interpreted by the clinical assessment of physicians, who followed the interim guidance for the clinical management of COVID-19 from the world health organization, May 27, 2020. 2) Patients with a SpO2 level less or equal to 90 %, exhibiting difficulty breathing, but not intubated patients. The physicians selected this set of patients because they had a low risk of developing a severe clinical condition with pneumonia or with a chance of being intubated. 3) Patients with similar pharmacological treatment against COVID-19. All the patients were medicated by physicians in respiratory medicine with Azithromycin (500 mg), Ivermectin (6 mg), and Oseltamivir (75 mg). The most prevalent comorbidities in these patients were diabetes and hypertension.

The exclusion criteria were as follows: 1) COVID-19 patients with acute respiratory failure requiring urgent intubation, 2) COVID-19 patients with impaired consciousness or during pregnancy, 3) patients with metallic implants in the thorax, abdomen or arms, or electronic medical devices such as pacemakers, and 4) children.

The criteria for discontinuing the LF-ThMS intervention were the participant request, SpO2 decrease, or any discomfort reported by the patient during the intervention. The strategy for achieving adequate participant enrolment to reach target sample size was the description of favorable results obtained from other patients

### Low-field thoracic magnetic stimulation (LF-ThMS) device

Dominguez-Nicolas developed the first custom-designed LF-ThMS device (patent pending by Dominguez-Nicolas SM, 2020) to modulate alternating current in a coil pair to generate low magnetic flux densities and magnetic hyperthermia for COVID-19 patients. Previous patents and experimental studies also induce magnetic hyperthermia [20-23], but they reach up to 71 °C (160 °F), not suitable for our application. Instead, we employed an electronic circuit in our LF-ThMS device to limit the temperature and magnetic flux density levels up to 44 °C and 13.1 mT for its safe use in the dorsal thorax of COVID-19 patients. In this form, we avoided harmfulness or adverse effects. The LF-ThMS device consisted of a virtual instrument, a PCI-DAS6031 acquisition board (Measurement Computing), an electronic board for the coupling between the digital signal and power, a power source of 0-15 Vcc, and 6-30 A, and two rings made of coils to generate LF-ThMS. We used 1.7 cm of cotton cloth disk between the LF-ThMS rings and the patient’s skin to allow homogeneous heat diffusion.

To generate the magnetic field, we used alternating current from 100 to 118 Hz with a peak amplitude of 8 A, polarized at 12 Vcc with a regulated power source of 0-15 Vcc and 6-30 A. We fabricated a couple of rings for LF-ThMS with an internal diameter of 9.5 cm and 130 turns. The LF-ThMS device emitted magnetic flux densities in the range of 10.5 to 13.1 mT. These were calculated theoretically with the Biot-Savart law and physically with a magnetic field sensor (475 DSP Gaussmeter, Lakeshore). We also used a thermocouple sensor (model NTC 10k) for monitoring temperature changes due to the LF-ThMS. Both the Gaussmeter and thermocouple sensors were helpful to calibrate magnetic flux densities and temperatures in a safe range (Table 1).

**Table 1.** Variables associated with the LF-ThMS (frequency, magnetic flux density, temperature) and the mean SpO2 levels in 17 COVD-19 patients.

| **Time**<br>**(min)** | **Frequency**<br>**(Hz)** | **Magnetic**<br>**flux density**<br>**(mT)** | **Temperature**<br>**°C** | **SpO2 (%)**<br>**mean±SD** | **Number**<br>**of**<br>**patients** |
| --- | --- | --- | --- | --- | --- |
| 0 | 0 | 0 | 27.5±0.2 | 86.6±2.2 | 17 |
| 5 | 100 | 10.5±0.01 | 29.3±0.07 | 86.7±2.0 | 17 |
| 10 | 103 | 10.7±0.04 | 35.2±0.08 | 88.0±2.1 | 17 |
| 15 | 105 | 11.1±0.12 | 41.4±0.3 | 88.8±2.2 | 17 |
| 20 | 110 | 11.6±0.02 | 43.3±0.2 | 90.1±2.4 | 17 |
| 25 | 115 | 12.2±0.2 | 44±0.05 | 91.4±2.5 | 17 |
| 30 | 118 | 13.1±0.15 | 43.9±0.01 | 92.2±3.2 | 17 |

The main electronic components of the LF-ThMS device consist of a power relay RL of two poles, a 12 Vcc coil, silver alloy contacts of Vcc/10A or 250 Vca/10A, Q NPN 2N2222 transistor, 10 kΩ resistance at 0.25 Watts, and a D IN4007 semiconductor diode. We also employed a PCI-DAS6031 board to energize the RL at 10 Vcc and a virtual instrument developed in Delphi Borland 7. Figure 1 shows the electronic circuit of our LF-ThMS device, and Table 1 shows the frequency, magnetic flux density, and temperature associated with the LF-ThMS during a 30 minutes single session.

**Figure 1.**
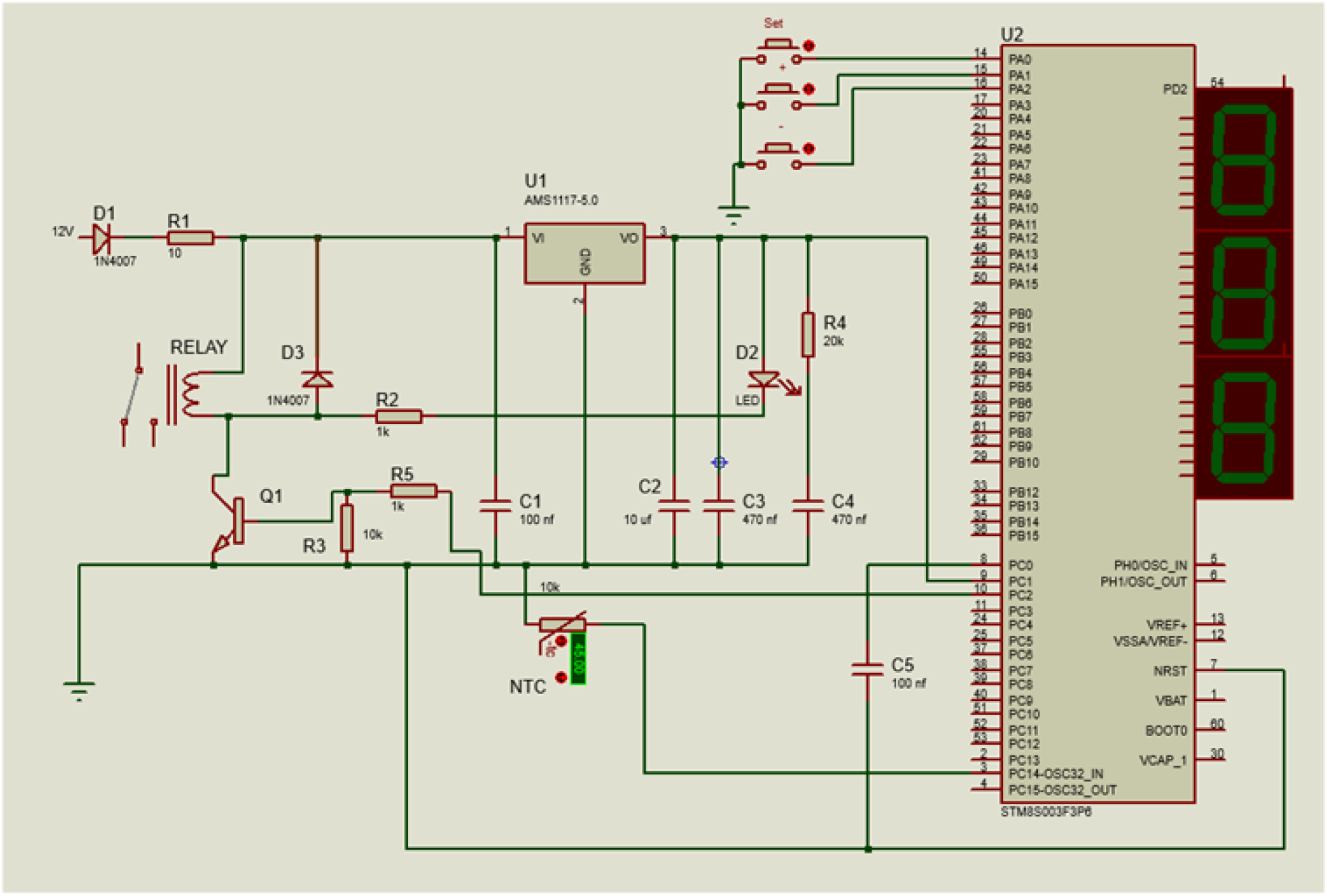
Electronic circuit employed in the low-field thoracic magnetic stimulation (LF-ThMS) device. This electronic circuit modulates the intensity and frequency of the alternating current producing the magnetic fields, limiting the maximum temperature to 44 °C, and the magnetic flux density up to 13.1 mT at 118 Hz.

### LF-ThMS protocol

The LF-ThMS was locally applied on the dorsal thorax while the patients were kept in a prone position. The LF-ThMS intensity was successively increased every 5 min during a single session of 30 min, following the values of frequency, magnetic flux density, and temperature indicated in Table 1. The protocol for the proof of concept consisted of a single LF-ThMS session of 30 minutes, although other daily sessions in three or four other consecutive days were applied to verify its reproducibility. The rationale of presenting here only the results from the first session was to evaluate the hypothesis that SpO2 levels in COVID-19 patients are significantly correlated with the magnetic flux density and temperature during 30 minutes of LF-ThMS intervention (see discussion section).

Figure 2A illustrates anatomical landmarks and coordinates of the LF-ThMS rings. We positioned the center of these LF-ThMS rings using palpable skeletal landmarks. We employed the spinous process of C7 (i.e., *vertebra prominens*) as zero landmarks (see a gray circle in Figure 2A). The center of these rings was positioned 8.5 cm below this landmark and bilaterally ± 6 cm on the dorsal thorax (see black arrows in Figure 2A).

**Figure 2.**
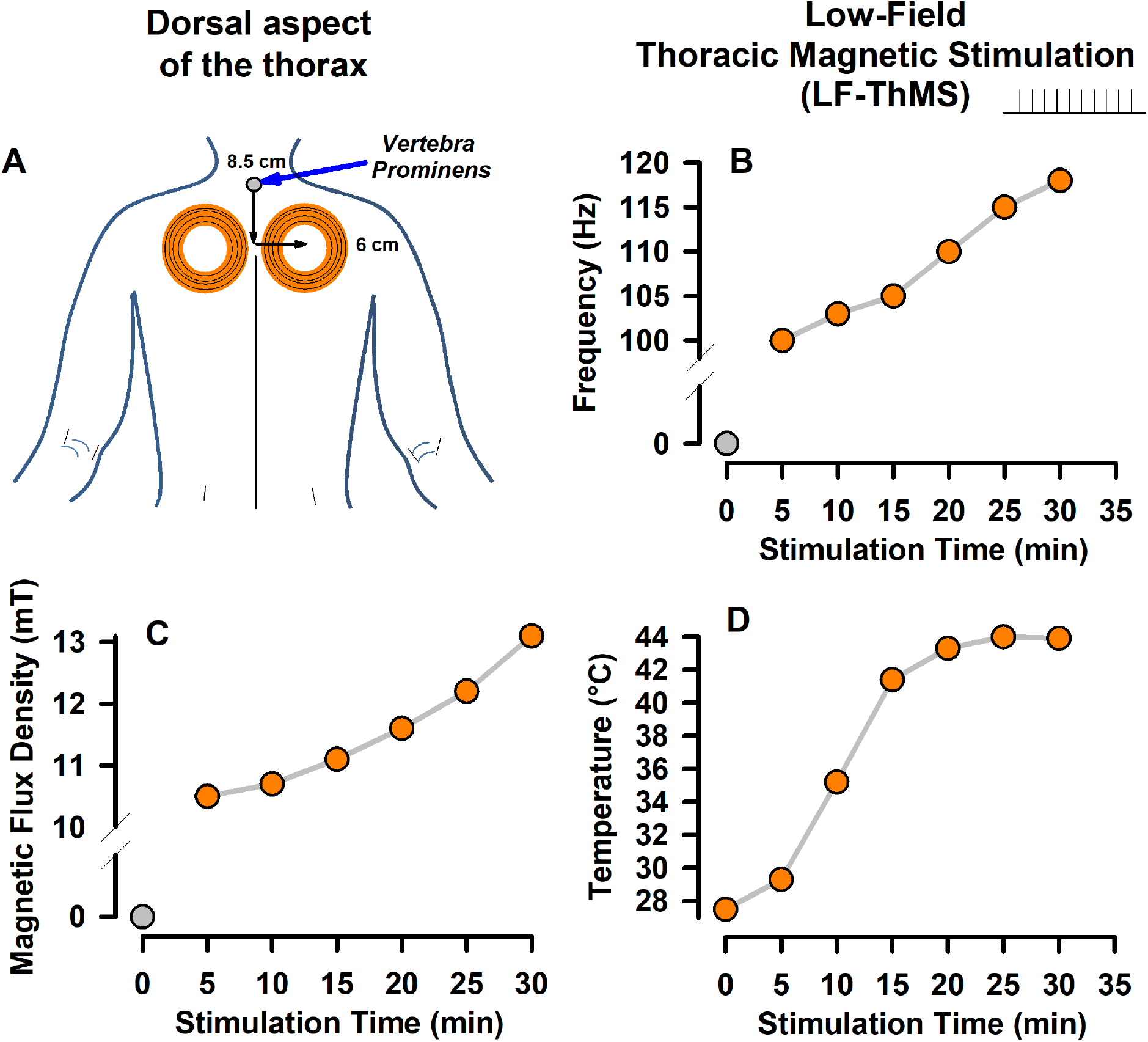
Experimental arrangement. **A**. Anatomical landmarks for the positioning of the LF-ThMS coils in the dorsal thorax of COVID-19 patients. The gray circle indicates the anatomical landmark called the “spinous process of C7” or “*vertebra prominens*”. **B-D**. Gradual increase (every 5 min) from 0 to 30 minutes of pulsed stimulus frequency, magnetic flux density, and temperature during the application of LF-ThMS to COVID-19 patients. The stimulation consisted of applying LF-ThMS for 30 minutes on the dorsal aspect of the thorax.

The device allowed a gradual increase in the frequency, magnetic flux density, and temperature, as illustrated in Figure 2B, 2C, and 2D, respectively. The patients rested for three hours after the session, and they did not report any discomfort during or after the magnetic stimuli. In contrast, they felt more comfortable, mainly because the LF-ThMS improved their breathing from the first minutes of the session. We checked on the health conditions of all the patients after receiving the LF-ThMS. In addition, in 11/17 patients, we monitored their SpO2 levels at the end of six months (see Results section).

### Sham stimulation

The coils were positioned in the same coordinates for sham exposure, but the pulse generator was not turned on. Subjects were blinded for the real LF-ThMS or sham stimulation conditions.

### Peripheral oxygen saturation (SpO2) level, magnetic flux density, and temperature monitoring

The peripheral oxygen saturation (SpO2) level was monitored with a conventional fingertip pulse oximeter (model C101H1) every 5 minutes during the 30 minutes LF-ThMS session. Thus, seven SpO2 measurements, including the control (time zero, at 27.5 °C and 0 mT, without LF-ThMS and compensating the terrestrial magnetic field), were obtained per subject. In this way, we were able to quantify the repeatability of effects of LF-ThMS through the study in different patients.

### Statistical analysis

We analyzed the statistical differences among the SpO2 levels related to each LF-ThMS intensity. For data normally distributed (Kolmogorov-Smirnov normality test, P>0.05) with homogeneity of variances, we used parametric one-way repeated-measures ANOVA under the null hypothesis that the dependent variables “SpO2 levels” were the same across the different LF-ThMS intensities. We also employed Mauchly’s test to verify that the assumption of sphericity was not violated. We performed a pairwise post hoc test using the corrected Bonferroni adjustment. All effects are reported as significant if p<0.001.

Moreover, a Pearson’s product-moment correlation coefficient was employed to examine whether there is a statistically significant linear correlation between SpO2 levels and frequency, magnetic flux density, and temperature changes during the LF-ThMS intervention. The sample size was n=35 SpO2 measurements during seven LF-ThMS levels (including the control) in five patients of the sham-controlled experiment and n=119 SpO2 values in another 12 patients. The correlation coefficient was calculated for n-2=33 or n-2=117 degrees of freedom (DF), and the correlation was reported as significant if p<0.001. Data were expressed as mean ± standard deviation in the main text and the figures.

## Results

We measured the SpO2 level for all the patients before the LF-ThMS intervention. We found that COVID-19 patients presented similar symptoms. On the first day of magnetic stimulation, we found that the patients experienced difficulty breathing with a low SpO2 level of 86.6 ± 2.2 % (N=17 patients), consistent with COVID-19 signs and breathlessness symptoms. However, we found that during the LF-ThMS, the patients exhibited a gradual increase in their SpO2 levels. No adverse events or discomfort sensations were reported during or after the LF-ThMS.

### Comparisons between the SpO2 levels of COVID-19 patients in the sham and real LF-ThMS

In the controlled study, we observed no statistically significant changes in SpO2 levels during sham stimulation (five subjects, Figure 3A). We performed parametric one-way repeated-measures ANOVA to examine statistical significance between SpO2 levels during the sham stimulation in all the patients. The differences in the mean values among the sham groups are not great enough to exclude the possibility that the difference is due to random sampling variability, i.e., there is no statistically significant difference [F=0.165, DF=6, p=0.984].

**Figure 3.**
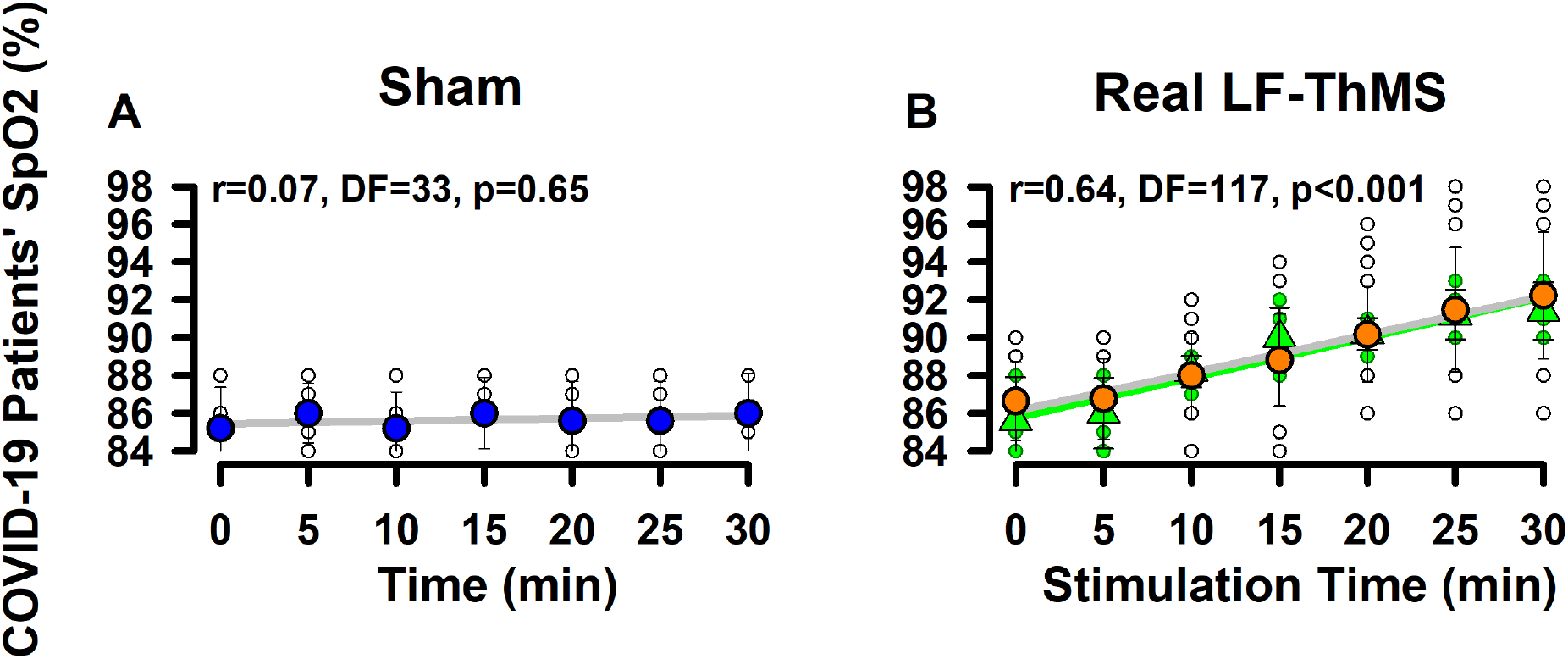
Comparative results obtained from the sham-controlled study and the real LF-ThMS intervention. **A**. Peripheral oxygen saturation (SpO2) levels versus time during the sham stimulation for five COVID-19 patients. Blue circles represent the grand average of these SpO2 levels **B**. SpO2 levels versus time during the LF-ThSM stimulation. The green triangles represent the grand average of SpO2 values during the real LF-ThSM in the same five patients that received sham stimulation in A. The orange symbols illustrate the grand average SpO2 levels in 12 COVID-19 patients versus time during a single session of LF-ThMS. The green and gray circles (raw data) show the SpO2 values obtained for all the patients (every 5 minutes) versus the time in minutes. A statistically significant correlation (p<0.001, Pearson’s product-moment correlation) was found for “SpO2 values” versus “stimulation time” during real LF-ThMS but not during sham stimulation.

However, during the real LF-ThMS, we observed statistically significant changes in SpO2 levels in response to real LF-ThMS in 5 COVID-19 subjects who previously underwent sham stimulation (green triangles and green line; Figure 3B) and in the other 12 COVID-19 subjects who underwent only the real LF-ThMS (orange circles and gray line; Figure 3B). We performed parametric one-way repeated-measures ANOVA to examine statistical significance between groups: “control SpO2 levels” and “SpO2 levels during the LF-ThMS interventions” in all the patients (17 subjects). The differences in the mean values among the treatment groups were more significant than would be expected by chance [F=13.872, DF=6, p<0.001]. The post hoc test indicated that the significant main effect exhibited significant differences (p<0.001) between the “control SpO2 levels” and the “SpO2 levels obtained after 20 min of LF-ThMS interventions” (Table 2). In contrast, no statistically significant differences (p>0.05) were found before 20 min of LF-ThMS interventions (Table 2). This indicates that the LF-ThMS at the frequency and magnetic flux density employed produces changes in SpO2 levels only after 20 minutes of LF-ThMS application.

**Table 2.** One way repeated measures ANOVA for the SpO2 values of 17 COVID-19 patients in control conditions (0 minutes) and during the LF-ThMS at 10, 15, 20, 25, and 30 minutes. The Bonferroni t-test was used for multiple comparisons versus the control group.

| Comparison | Diff of Means | t | P | Significance |
| --- | --- | --- | --- | --- |
| SpO2 (0 min) vs. SpO2 (30 min) | 5.588 | 6.655 | <0.001 | Yes |
| SpO2 (0 min) vs. SpO2 (25 min) | 4.824 | 5.745 | <0.001 | Yes |
| SpO2 (0 min) vs. SpO2 (20 min) | 3.529 | 4.203 | <0.001 | Yes |
| SpO2 (0 min) vs. SpO2 (15 min) | 2.176 | 2.592 | 0.065 | No |
| SpO2 (0 min) vs. SpO2 (10 min) | 1.353 | 1.611 | 0.66 | No |
| SpO2 (0 min) vs. SpO2 (5 min) | 0.118 | 0.14 | 1 | No |

### Correlations among the frequency, magnetic flux density, and temperature elicited by the LF-ThMS versus the SpO2 levels of COVID-19 patients

In the controlled study, we also examined whether SpO2 levels exhibited correlations with the sham session time. We observed no statistically significant correlation between SpO2 levels and session time in the sham condition (Pearson’s product-moment correlation coefficient r=0.07, DF=33, p=0.65, gray regression line, Figure 3A). However, we obtained a statistically significant correlation between SpO2 values and session time in the real LF-ThMS applied to the same five subjects that previously underwent the sham stimulation (Pearson’s product-moment correlation coefficient r=0.81, DF=33, p<0.001, green regression line, Figure 3B).

Finally, we examined whether the SpO2 levels exhibited correlations with the LF-ThMS session time, frequency, magnetic flux density, and temperature for all the patients. Figure 3B and Figures 4A, 4B, and 4C show significant correlations between these measurements. Pearson’s product-moment correlation method was used to test for significant correlations. We obtained a p<0.001 with 117 or 100 degrees of freedom and correlation coefficients r = 0.64, 0.58, 0.55, and 0.64, as indicated (see Figure 3B, and Figures 4A, 4B, and 4C). These statistically significant results suggest that the changes in SpO2 levels during a 30 min LF-ThMS session are related to their associated variables: frequency, magnetic flux density, and temperature. These findings provide support to our hypothesis.

**Figure 4.**
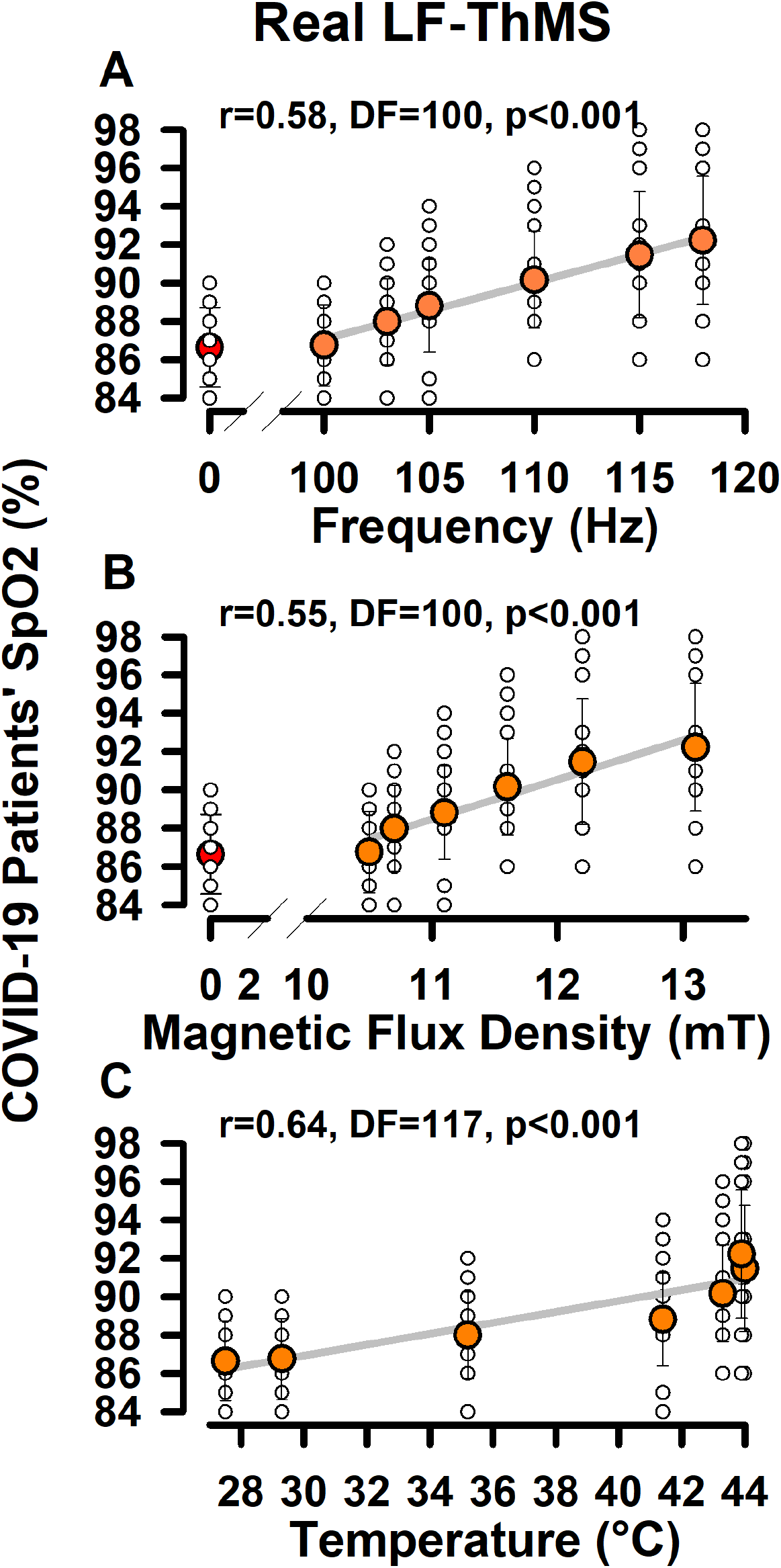
The same as Figure 3B, but for the correlation between SpO2 levels and the variables related to the real LF-ThMS applied to 17 COVID-19 patients. The Pearson’s correlation coefficients (r), degrees of freedom (DF), and p values (p<0.001) are shown above each graph.

### Adverse events

We did not find adverse effects during or after the LF-ThMS intervention. We also made a follow-up on the health conditions of all the patients. We found that five days after successive LF-ThMS sessions, the mean SpO2 level was 98.3 ± 0.7 % for 17 patients (Table 3). We also found a mean SpO2 level of 98.4 ± 0.8 %, for 11 patients, six months after the LF-ThMS intervention (Table 3). Such normal SpO2 levels indicate that the LF-ThMS did not produce adverse events in the oxygen saturation. In the follow-up on the general health conditions after five days or six months, the physicians confirmed that the patients did not exhibit any adverse event or secondary effect after the LF-ThMS intervention. These findings indicate that 30 minutes of LF-ThMS intervention on the dorsal thorax of COVID-19 patients is safe at the frequencies, magnetic flux densities, and temperatures of 100 to 128 Hz, 10.5 to 13.1 mT, and 27.5 to 44 °C, respectively.

**Table 3.** Follow-up on the SpO2 values for all the patients after the LF-ThMS. Six months after the LF-ThMS intervention, the patients reported no adverse events, and they exhibited normal SpO2 levels.

|  | Before<br>LF-ThMS | 30 minutes after<br>Sham Stimulus | 30 minutes after<br>LF-ThMS | Five days after<br>LF-ThMS | Six months after<br>LF-ThMS |
| --- | --- | --- | --- | --- | --- |
| **Patients** | **SpO2 (%)** | **SpO2 (%)** | **SpO2 (%)** | **SpO2 (%)** | **SpO2 (%)** |
| Patient 1 | 88 | — | 98 | 98 | 99 |
| Patient 2 | 86 | — | 97 | 98 | 97 |
| Patient 3 | 89 | — | 96 | 98 | 97 |
| Patient 4 | 90 | — | 97 | 99 | 99 |
| Patient 5 | 87 | — | 91 | 98 | 99 |
| Patient 6 | 90 | — | 93 | 99 | 99 |
| Patient 7 | 87 | — | 91 | 97 | 98 |
| Patient 8 | 88 | — | 93 | 98 | 98 |
| Patient 9 | 87 | — | 91 | 99 | 99 |
| Patient 10 | 86 | — | 90 | 98 | 99 |
| Patient 11 | 84 | — | 88 | 97 | 99 |
| Patient 12 | 83 | — | 86 | 99 | — |
| Patient 13 | 84 | 85 | 93 | 99 | — |
| Patient 14 | 85 | 86 | 90 | 99 | — |
| Patient 15 | 88 | 88 | 93 | 99 | — |
| Patient 16 | 83 | 83 | 90 | 98 | — |
| Patient 17 | 88 | 88 | 91 | 99 | — |
|  | 86.6 ± 2.2 | 86 ± 2.1 | 92.2 ± 3.2 | 98.3 ± 0.7 | 98.4 ± 0.8 |

## Discussion

We found statistically significant correlations between SpO2 levels in COVID-19 patients and LF-ThMS variables in a time range of 30 minutes, but not in the sham-controlled study.

### Reproducibility

Our findings were reproducible in all the patients in a time range of 30 minutes of LF-ThMS intervention. Interestingly, our results were also reproducible in three or four other subsequent sessions. However, we did not present such data because the changes could be associated with an ongoing daily recovery of the patients due to unknown immune mechanisms and not necessarily due to the daily LF-ThMS intervention. Hence, future studies of randomized controlled trials will be necessary to examine the potential use of this LF-ThMS application as therapy during consecutive daily sessions in covid-19 patients. Therefore, the principal value of our results is that in the sham-controlled study, we found a reproducible and significant correlation between LF-ThMS associated variables and SpO2 levels in a short time range of 30 minutes.

### Possible interference of LF-ThMS hyperthermia with the virus-host protein interactions

It is well known that several viral protein complexes mediate the entry and replication of SARS-CoV-2 into the cells, manipulating the host mRNA translation, subsequent viral protein production, antivirus immunity, and inflammation response to inducing lung infection and pneumonia. Specifically, this pathogen is a single-stranded RNA virus with gene fragments expressing structural and nonstructural proteins [24-26].

Several viral protein complexes are involved in the entry and replication of this virus into the cells; for instance, the virus spike (S) protein and the nonstructural protein 1 (nsp1). The first one mediates cell entry via binding with angiotensin-converting enzyme 2 in host cells (AEC2), and the second one, the crucial virus-host interactions [27,28]. Because there is evidence that a temperature increase of tissue can affect proteins and enhance the immune response [14,15], it is tempting to speculate that hyperthermia produced by the LF-ThMS may acutely interfere with these viral proteins and improve respiratory function.

### Possible interference of LF-ThMS magnetic flux with the virus-host electrical interactions

Another possibility is that the magnetic stimuli could also directly interfere with the positively charged site in the SARS-CoV-2 spike protein, disturbing the electrical binding between the virus protein and the negatively charged human-cell receptors. This is consistent with recent simulation studies, reporting a positively charged site (called polybasic cleavage site) positioned 10 nanometers from the actual binding site on the SARS-CoV-2 spike protein [29]. These authors found that the positively charged site allows strong bonding between the virus protein and the negatively charged cell receptors. In their simulation, Qiao and Olvera de la Cruz [29] designed a negatively charged molecule to bind to the positively charged cleavage site, with the idea that blocking this site inhibits the virus from bonding to the host cell [29]. Therefore, it is also tempting to speculate that interfering with the electrostatic interaction during the binding action of the SARS-CoV-2 spike protein and the ACE2 receptors, or the nsp1 could mitigate the viral infection. This possible mode of action of the LF-ThMS is also consistent with the claim that electrostatic precipitators are also valuable for eliminating airborne virus particles [30,31].

### Possible interference of LF-ThMS hyperthermia with the immune response and interferon activity

Another mechanism by which the LF-ThMS up to 44 °C could improve SpO2 levels in COVID-19 patients is the enhanced immune response due to the dorsal thorax’s increased temperature during the intervention. This is consistent with reports that hyperthermia potentiates the immune response against cancer through immune cells’ activation [15,32,33]. Some of the immune cells activated by hyperthermia are the natural killer cells, dendritic cells, and cytotoxic T-lymphocytes, which alter the cell-surface molecules on cancer cells, modifying adhesion molecules on immune cells and endothelial cells [32].

Previous studies claimed that Interferons could have a potential role in treating COVID-19 patients [34]. However, recent investigations demonstrated that the SARS-CoV-2 receptor ACE2 is an Interferon-stimulated gene in human airway epithelial cells [35]. This means that interferons could help or damage, depending on the infection stage for each COVID-19 patient. In this context, we suggest that LF-ThMS should be applied at the first stages of the COVID-19 infection to enhance interferon activity by an increased temperature, in which the interferons may confer an antiviral state on cells.

### Possible interference of LF-ThMS in inflammation and cytokine storm

Finally, because the inflammation and cytokine storm are the main factors contributing to breathing, ventilation, and oxygenation, in COVID-19 patients, it will be necessary to examine in future studies whether the LF-ThMS has an impact on these factors.

### Advantages and limitations

The first potential advantage of the thoracic LF-ThMS is that the subjects did not require oxygen therapy with face masks, mainly because during the LF-ThMS session, the patients significantly increased their SpO2 levels 20 minutes after the LF-ThMS (p<0.001, Table 2). The second advantage is that the device for LF-ThMS is easy to reproduce, and the electronic components are not expensive. It may be possible that several pulsed electromagnetic field devices employed in physical therapy worldwide could be adapted to emit magnetic fields at 100 to 118 Hz, 10.5 to 13.1 mT (105 to 131 Gauss), and 27.5 to 44 °C.

The main limitations of our study are the following. We do not know the physiological mechanisms through which the administered LF-ThMS during a 30 min session improved SpO2 levels in COVID-19 patients. We also did not explore whether the LF-ThMS intervention enhances the patients’ immune response or whether it impacts the electrical charges of the SARS-CoV-2 viral proteins or the inflammation and cytokine storm in COVID-19 patients. However, our study will motivate future investigations in this research field.

Another limitation of our study is that it will be necessary to know the real temperature in the lungs associated with variations in temperature of the external dorsal thorax by LF-ThMS. It is expected that such temperatures should be lower than those on the external dorsal thorax due to the diffusion processes of heat transfer occurring in the skin, muscle, and scapula. In the same context, it will be necessary to examine the magnetic flux density reached into the lungs, which should be attenuated as a function of depth.

Finally, while there is a consensus that repetitive magnetic stimulation is helpful in the non-invasive modulation of brain neural activity in humans, the use of similar interventions in other regions of the human body is still controversial. Besides such a limitation, here we found the experimental correlation of LF-ThMS variables: magnetic flux density, frequency, and temperature with SpO2 levels in 17 COVID-19 patients, five of them in a single-blind, sham-controlled, crossover study.

### Perspectives

Although the increased SpO2 levels may be attributed to the altered perfusion consequent to warming or the impact of magnetic factors inactivating the SARS-CoV-2 virus, at this moment, these are only speculations. Therefore, future studies will be necessary to examine the physiological mechanisms of these significant correlations.

Besides previous studies suggest that magnetic stimulation could be helpful in chronic obstructive pulmonary disease [36] and phrenic nerve activation [37], future studies examining this issue by using the LF-ThMS in humans or animal preparations will also be necessary. Other future perspectives include developing wearable and portable devices for LF-ThMS, combined with oximeters and respiratory magnetograms [38]. Such devices could help examine in more detail respiratory improvements after the thoracic LF-ThMS in COVID-19 patients.

Here our LF-ThMS protocol is not intended to demonstrate its use as therapy but is instead designed to examine the hypothesis that LF-ThMS could be helpful to increase SpO2 levels in COVID-19 patients in a short range from 0 to 30 minutes. In this context, our findings are relevant because they could motivate future randomized clinical trials to examine whether the LF-ThMS could be helpful as a potential therapy.

## Conclusions

We conclude that our findings are relevant at this stage, mainly because they provide evidence that 30 minutes of LF-ThMS on the dorsal thorax at 100 to 118 Hz, 10.5 to 13.1 mT (105 to 131 Gauss), and 27.5 to 44 °C is safe in COVID-19 patients. We also conclude that the LF-ThMS variables (frequency, magnetic flux density, and temperature) exhibit a statistically significant correlation with SpO2 levels in the time range of 30 minutes, showing that the LF-ThMS significantly increases peripheral oxygen saturation levels in COVID-19 patients.

## Supporting information

SPIRIT-Checklist

## Data Availability

The authors confirm that all data underlying the findings are fully available without restriction upon request. All relevant data are within the paper.

## Ethics and reporting

We declare that all relevant ethical guidelines have been followed and ethics committee approval has been obtained. All necessary patient/participant consent has been obtained. SPIRIT-Checklist was included as a supplementary file.

## Acknowledgments

We thank Prof. Robert Simpson for proofreading the English document.

## Author contributions

SMDN conceived the therapeutic application of magnetic stimuli in COVID-19 patients, developed the first LF-ThMS prototype, and performed the experiments. EM and SMDN conceived the proof-of-concept and hypothesis of this study. EM performed the data analysis. EM designed the single-blind, sham-controlled, crossover study on 5 COVID-19 patients, wrote the paper, and contributed to calibrate and improve the safeness of the original LF-ThMS prototype.

