## Supplementary material for "Low-field thoracic magnetic stimulation increases peripheral oxygen saturation levels in coronavirus disease (COVID-19) patients: a single-blind, sham-controlled, crossover study": SPIRIT-Checklist

SPIRIT
STANDARD PROTOCOL ITEMS: RECOMMENDATIONS FOR INTERVENTIONAL TRIALS

SPIRIT 2013 Checklist: Recommended items to address in a clinical trial protocol and related documents*

| Section/item | Item No | Description |
|---|---|---|
| **Administrative information** | | |
| Title | 1 | Low-field Thoracic Magnetic Stimulation Increases Peripheral Oxygen Saturation Levels in Coronavirus Disease (**COVID-19**) Patients: a Single-blind, Sham-controlled, Crossover Study |
| Trial registration | 2a | ClinicalTrials.gov Identifier: NCT04895267<br>Low-field Thoracic Magnetic Stimulation Increases Peripheral Oxygen Saturation Levels in COVID-19 Patients<br>https://clinicaltrials.gov/ct2/show/NCT04895267?cond=Covid19&cntry=MX&city=Puebla&draw=2&rank=3 |
| Protocol version | 3 | First Posted: May 20, 2021<br>Protocol version 1 |
| Funding | 4 | Centro de Investigación de Micro y Nanotecnología (SMDN), National Council of Science and Technology, Mexico, CONACyT Fronteras de la Ciencia #536 (EM), Cátedra Marcos Moshinsky (EM), and VIEP-PIFI-FOMES-PROMEP-BUAP-Puebla (EM), Comité de Internacionalización and VIEP BUAP (EM), México. |
| Roles and responsibilities | 5a | The principal investigators share all the responsabilities:<br>Dr. Saul M Dominguez-Nicolas<br>Centro de Investigación de Micro y Nanotecnología, Universidad Veracruzana<br>Calzada Ruiz Cortines 455 Boca del Rio, Veracruz 94294, Mexico<br><br><br>Dr. Elias Manjarrez<br>Instituto de Fisiología, Benemérita Universidad Autónoma de Puebla.<br>14 sur 6301, Col. San Manuel A.P. 406, C.P. 72570<br>Puebla, Pue., México<br>Tels.: +5222-22-29-5500 Ext 7326, Fax: +5222-22-33-4511<br> |
| | 5b | No applicable |

**Methods: Participants, interventions, and outcomes**

| | | |
|---|---|---|
| Study setting | 9 | No applicable for MedRxiv |

| | | |
|---|---|---|
| Eligibility criteria | 10 | The inclusion criteria were: 1) Adult patients diagnosed with mild to moderate COVID-19 disease without pneumonia. The COVID-19 disease severity was interpreted by the clinical assessment of physicians from the Mexican Institute of social security or physicians from private hospitals, who followed the interim guidance for the clinical management of COVID-19 from the world health organization 27 May 2020. 2) Patients with a SpO2 level less or equal to 90 %, exhibiting difficulty breathing, but not intubated patients. The physicians selected this set of patients because they had a low risk of developing a severe clinical condition with pneumonia or with a chance of being intubated. 3) Patients with the same pharmacological treatment against COVID-19. All the patients were medicated with Azithromycin (500 mg), Ivermectin (6 mg), and Oseltamivir (75 mg) by physicians from the health sector. The most prevalent comorbidities in these patients were diabetes and hypertension.<br>The exclusion criteria were as follows: 1) COVID-19 patients with acute respiratory failure requiring intubation, 2) COVID-19 patients with impaired consciousness or during pregnancy, 3) patients with metallic implants or electronic medical devices such as pacemakers, and 4) children. |
| Interventions | 11a | The low field thoracic magnetic stimulation (LF-ThMS) was locally applied on the dorsal thorax while the patients were kept in a prone position. The protocol for the proof of concept consisted of a single LF-ThMS session of 30 minutes. The rationale of presenting the LF-ThMS was to evaluate the hypothesis that SpO2 levels in COVID-19 patients are significantly correlated with the magnetic flux density and temperature during 30 minutes of LF-ThMS intervention.<br>We positioned the center of these LF-ThMS rings using palpable skeletal landmarks. We employed the spinous process of C7 (i.e., vertebra prominens) as zero landmarks. The center of these rings was positioned 8.5 cm below this landmark and bilaterally ± 6 cm on the dorsal thorax (see black arrows in Figure 2A).<br>The device allowed a gradual increase in the frequency, magnetic flux density, and temperature. The patients rested for three hours after the session, and they did not report any discomfort during or after the magnetic stimuli. In contrast, they felt more comfortable, mainly because the LF-ThMS improved their breathing from the first minutes of the session. We checked on the health conditions of all the patients after receiving the LF-ThMS. In addition, in 11/17 patients, we monitored their SpO2 levels at the end of six months.<br>For sham exposure, the coils were positioned in the same coordinates, but the pulse generator was not turned on. Subjects were blinded for the real LF-ThMS or sham stimulation conditions. |
| | 11b | The criteria for discontinuing the LF-ThMS intervention were the participant request, SpO2 decrease or any discomfort reported by the patient. |

| | | |
|---|---|---|
| | 11c | No applicable |
| | 11d | No concomitant interventions were applied during the trial |
| Outcomes | 12 | We made a follow-up on the health conditions of all the patients after the LF-ThMS intervention up to 6 months. SpO2 levels were monitored five days after the intervention, and at the end of 6 months. |
| Participant timeline | 13 | A single intervention of 30 minutes per patient was obtained. |
| Sample size | 14 | The estimated number of participants needed to achieve study objectives was 17 and 5 patients for the sham-controlled trial. This sample size was determined from the statistical significant differences reached at $p<0.001$. |
| Recruitment | 15 | The strategy for achieving adequate participant enrolment to reach target sample size was the description of favourable results obtained from other patients. |

**Methods: Assignment of interventions (for controlled trials)**

Allocation:

| | | |
|---|---|---|
| Sequence generation | 16a | There was not an allocation sequence for these patients |
| Allocation concealment mechanism | 16b | The interventions were assigned by a request of the patients |
| Implementation | 16c | No applicable |
| Blinding (masking) | 17a | For sham exposure, the coil was positioned in the same coordinates, but the pulse generator was not turned on. Subjects were blinded for the real LF-ThMS or sham stimulation conditions. |
| | 17b | Unblinding was not permissible during the single-blind, sham-controlled trial. |

**Methods: Data collection, management, and analysis**

| | | |
|---|---|---|
| Data collection methods | 18a | Data were collected for statistical analysis and questionnaires were employed for a follow-up on the health conditions of all the patients after the LF-ThMS intervention. |
| | 18b | All patients had a follow-up |
| Data management | 19 | No applicable |

| | | |
|---|---|---|
| Statistical methods | 20a | We analyzed the statistical differences among the SpO2 levels related to each LF-ThMS intensity. For data normally distributed (Kolmogorov-Smirnov normality test, P>0.05) with homogeneity of variances, we used parametric one-way repeated-measures ANOVA under the null hypothesis that the dependent variables "SpO2 levels" were the same across the different LF-ThMS intensities. We also employed Mauchly's test to verify that the assumption of sphericity was not violated. We performed a pairwise post hoc test using the corrected Bonferroni adjustment. All effects are reported as significant if p<0.001.<br>Moreover, a Pearson's product-moment correlation coefficient was employed to examine whether there is a statistically significant linear correlation between SpO2 levels and frequency, magnetic flux density, and temperature changes during the LF-ThMS intervention. The sample size was n=35 SpO2 measurements during seven LF-ThMS levels (including the control) in five patients of the sham-controlled experiment and n=119 SpO2 values in another 12 patients. The correlation coefficient was calculated for n-2=33 or n-2=117 degrees of freedom (DF), and the correlation was reported as significant if p<0.001. Data were expressed as mean ± standard deviation in the main text and the figures. |
| | 20b | No applicable |
| | 20c | No applicable |
| **Methods: Monitoring** | | |
| Data monitoring | 21a | The data monitoring committee (DMC) was not needed given the low number of patients and the type of data generated. |
| | 21b | No applicable |
| Harms | 22 | Researchers participating in the study directly payed special attention in possible adverse events. |
| Auditing | 23 | No applicable |
| **Ethics and dissemination** | | |
| Research ethics approval | 24 | Researchers participating in the study directly payed special attention in the ethics guidelines |
| Protocol amendments | 25 | No applicable |
| Consent or assent | 26a | The patients signed an informed consent |
| | 26b | No applicable |

| | | |
|---|---|---|
| Confidentiality | 27 | The personal information about enrolled participants was confidentially collected, before, during, and after the trial |
| Declaration of interests | 28 | The principal investigators declare that there are not financial or other competing interests for the trial and study |
| Access to data | 29 | The authors confirm that all data underlying the findings are fully available without restriction upon request. All relevant data are within the paper. |
| Ancillary and post-trial care | 30 | No applicable |
| Dissemination policy | 31a | There are not publication restrictions |
| | 31b | No applicable |
| | 31c | The principal investigators grant public access to the full protocol, participant-level dataset, and statistical code |
| **Appendices** | | |
| Informed consent materials | 32 | No applicable for MedRxiv |
| Biological specimens | 33 | No applicable |

*It is strongly recommended that this checklist be read in conjunction with the SPIRIT 2013 Explanation & Elaboration for important clarification on the items. Amendments to the protocol should be tracked and dated. The SPIRIT checklist is copyrighted by the SPIRIT Group under the Creative Commons "Attribution-NonCommercial-NoDerivs 3.0 Unported" license.
